# Sleep Abnormalities in the Synaptopathies – *SYNGAP1*-related Intellectual Disability and Phelan-McDermid syndrome

**DOI:** 10.1101/2020.08.04.20168286

**Authors:** Constance Smith-Hicks, Robert C. Stowe, Maria McCormack, J. Lloyd Holder

**Affiliations:** Division of Neurogenetics Kennedy Krieger Institute, Baltimore, MD; Department of Neurology, Johns Hopkins University School of Medicine, Baltimore MD; Boston Children’s Hospital, Boston, MA; Jan and Dan Duncan Neurological Research Institute at Texas Children’s Hospital; Departments of Pediatrics and Neurology, Baylor College of Medicine, Houston, TX

**Keywords:** Phelan-McDermid syndrome, *SYNGAP1*, Children’s Sleep Habits Questionnaire, Polysomnography

## Abstract

Neurodevelopmental disorders are frequently associated with sleep disturbances. One class of neurodevelopmental disorders, the genetic synaptopathies, is caused by mutations in genes encoding proteins found at the synapse. Mutations in these genes cause derangement of synapse development and function. We utilized a validated sleep instrument, Children’s Sleep Habits Questionnaire (CSHQ) to discover what sleep abnormalities occur in individuals with two synaptopathies – Phelan-McDermid syndrome (PMD) and *SYNGAP1*-related Intellectual Disability (*SYNGAP1*-ID) when compared with healthy controls. We found both PMD and *SYNGAP1*-ID have significant sleep abnormalities with *SYNGAP1*-ID having greater severity of sleep disturbance than PMD. We found that sleep disturbances were more severe for both disorders in individuals 10 years and older compared with those less than 10 years old. Individuals with either disorder were more likely to use sleep aids than healthy controls.

Furthermore, review of polysomnography studies for individuals with *SYNGAP1*-ID revealed significant reduction in rapid eye movement (REM) sleep content and delayed REM latency demonstrating abnormalities in sleep architecture. In conclusion, sleep disturbances are a significant phenotype in the synaptopathies PMD and *SYNGAP1*-ID. Improved sleep is a viable clinical endpoint for future clinical trials for these neurodevelopmental disorders.

## INTRODUCTION

Neurodevelopmental disorders (NDDs) are severe clinical consequences of abnormal brain development. The phenotypic manifestations of NDDs include learning disabilities, delayed ascertainment of developmental milestones, intellectual disabilities, behavioral disturbances and epilepsy among others. NDDs can have multiple etiologies including sequelae of infections of the central nervous system either *in utero* or perinatally, restricted blood flow and oxygenation due to perinatal stroke or perinatal head trauma. Increasingly, genetic abnormalities are recognized as a significant cause of neurodevelopmental disorders. These genetic abnormalities include chromosomal derangements such as Trisomy 21, contiguous gene deletion syndromes or single gene disruptions.

Among all genetic NDDs, synaptopathies are unique in that they are caused by mutations in genes which encode proteins that function at the synapse^1^. The result of these mutations are dysfunctional synapses or alterations in synapse numbers. Both of these result in abnormalities in neuronal network development and consequent neuropsychiatric phenotypes. Two synaptopathies, Phelan-McDermid syndrome (PMD) and *SYNGAP1*-related intellectual disability (*SYNGAP1*-ID) have characteristic neurodevelopmental deficits including intellectual disability, global developmental delay, autism and epilepsy^2-6^.

Phelan-McDermid syndrome most commonly results from a deletion of chromosome 22q13 encompassing the *SHANK3* gene and variably other genes^7^. Single nucleotide variants in *SHANK3* have also been identified to cause PMD^5^. SHANK3 is a scaffolding protein of the post-synaptic density of excitatory synapses. *SYNGAP1*-related Intellectual Disability results most commonly from single nucleotide variants that cause loss-of-function mutations^4,6^. The SynGAP protein is enriched in the post-synaptic density where it functions as a small GTPase activating protein^8^.

Sleep abnormalities are commonly identified in children with NDDs in general and specifically in children with synaptopathies^4,9,10^. The pathophysiology of sleep disturbances in NDDs is multifactorial due to the interplay of genetic, neurobiological and environmental factors. Sleep abnormalities manifest as insomnia, hypersomnia, parasomnias or circadian dysregulation. Sleep disorders in children with NDDs commonly last into adolescence and adulthood^11^. For both PMD and *SYNGAP1*-ID, several reports have described sleep abnormalities^4-7^. However, quantitative measures of sleep disturbance have not been systematically applied to *SYNGAP1*-ID.

The primary goal of this study was to quantify, compare and contrast sleep disturbances in individuals with two synaptopathies, PMD and *SYNGAP1*-ID. We utilized a standardized instrument, the Children’s Sleep Habits Questionnaire (CSHQ), in order to identify sleep abnormalities in these two disorders.

## METHODS

### Recruitment

Participants for this study were recruited from the Bluebird Circle Clinic for Pediatric Neurology at Texas Children’s Hospital and the Kennedy Krieger Institute with the assistance of family advocacy foundations: Phelan McDermid Syndrome Foundation, Bridge the Gap: *SYNGAP1* Education and Research Foundation and the SynGap Research Fund, Inc. We provided the patient advocacy organizations with a recruitment letter that was sent by email to their registrants. Adult caregivers of individuals from age 12 months and above with Phelan-McDermid syndrome and *SYNGAP1*-related intellectual disability were eligible to participate. Those interested in participating contacted the research staff and informed consent was obtained. The research staff either directly asked the caregivers 33 total questions on the Children’s Sleep Habit Questionnaire (CSHQ) or the caregivers completed an online version of the CSHQ. This instrument assesses parental reports of frequency of various sleep behaviors during a typical week as ‘usually’ (5-7 times/week),‘sometimes’ (2-4 times/week) or ‘rarely’ (0-1 times/week); higher scores indicate worse sleep problems or behaviors. Answers to the questionnaire were recorded once and was completed in 15-20 minutes. The caregivers were also asked for typically developed siblings to participate as healthy control individuals. The surveys were performed between May 2019 to June 2020. Ethical approval was obtained from the Institutional Review Board for Baylor College of Medicine and Affiliated Hospitals and the Johns Hopkins University School of Medicine.

### Data and Statistical Analysis

Raw data were aggregated in Microsoft Excel. These data were then both summed to determine the total score per individual or parsed into eight sub-scales: bedtime resistance, sleep anxiety, sleep-onset delay, night awakenings, parasomnias, daytime sleepiness, sleep disordered breathing and sleep duration for calculating sub-scale scores per each individual. Statistical analysis for comparison of PMD, *SYNGAP1*-ID and healthy control data was performed in GraphPad Prism version 8. For comparing all three groups, Kruskal-Wallis non-parametric tests were used. For direct comparison of either *SYNGAP1*-ID or PMD versus healthy controls, Dunn’s multiple comparison tests were performed.

## RESULTS

The participants’ average age was not significantly different between *SYNGAP1*-ID (7.6±4.5 years; Dunn’s multiple comparison’s test p=.77) or PMD (12.6±9.6 years; Dunn’s multiple comparison’s test p=.26) and healthy controls (8.7±4.8 years). For *SYNGAP1*-ID, we recruited 17 males (52%) and 16 females (48%). For PMD, 17 males (44%) and 22 females (56%) were recruited. These synaptopathy subjects were compared with 14 male (37%) and 24 female (63%) healthy controls who were all siblings of synaptopathy subjects.

The CSHQ average total score (Figure 1 and Table I) for *SYNGAP1*-ID (M=54.5±8.3) was significantly increased (p<0.0001) compared with healthy controls (M=44.3±8.1). In contrast, there was a smaller average increased total score (M=48.4±8.6) for PMD that did not reach statistical significance (p=.08). We then evaluated sub-scale scores between *SYNGAP1*-ID or PMD and healthy controls. Subjects with *SYNGAP1*-ID had significantly increased bedtime resistance, sleep anxiety and sleep duration compared with healthy controls (Figure 1 and Table I). No significant change was seen in these sub-scales for PMD. For sleep onset delay, night-time awakenings, parasomnias and sleep disordered breathing, we found significantly elevated scores in both *SYNGAP1*-ID and PMD compared with healthy controls (Figure 1 and Table I). Only for daytime sleepiness did we find a significant difference in only PMD and not *SYNGAP1*-ID, and in this case PMD patients scored lower than healthy controls.

**Figure 1.**
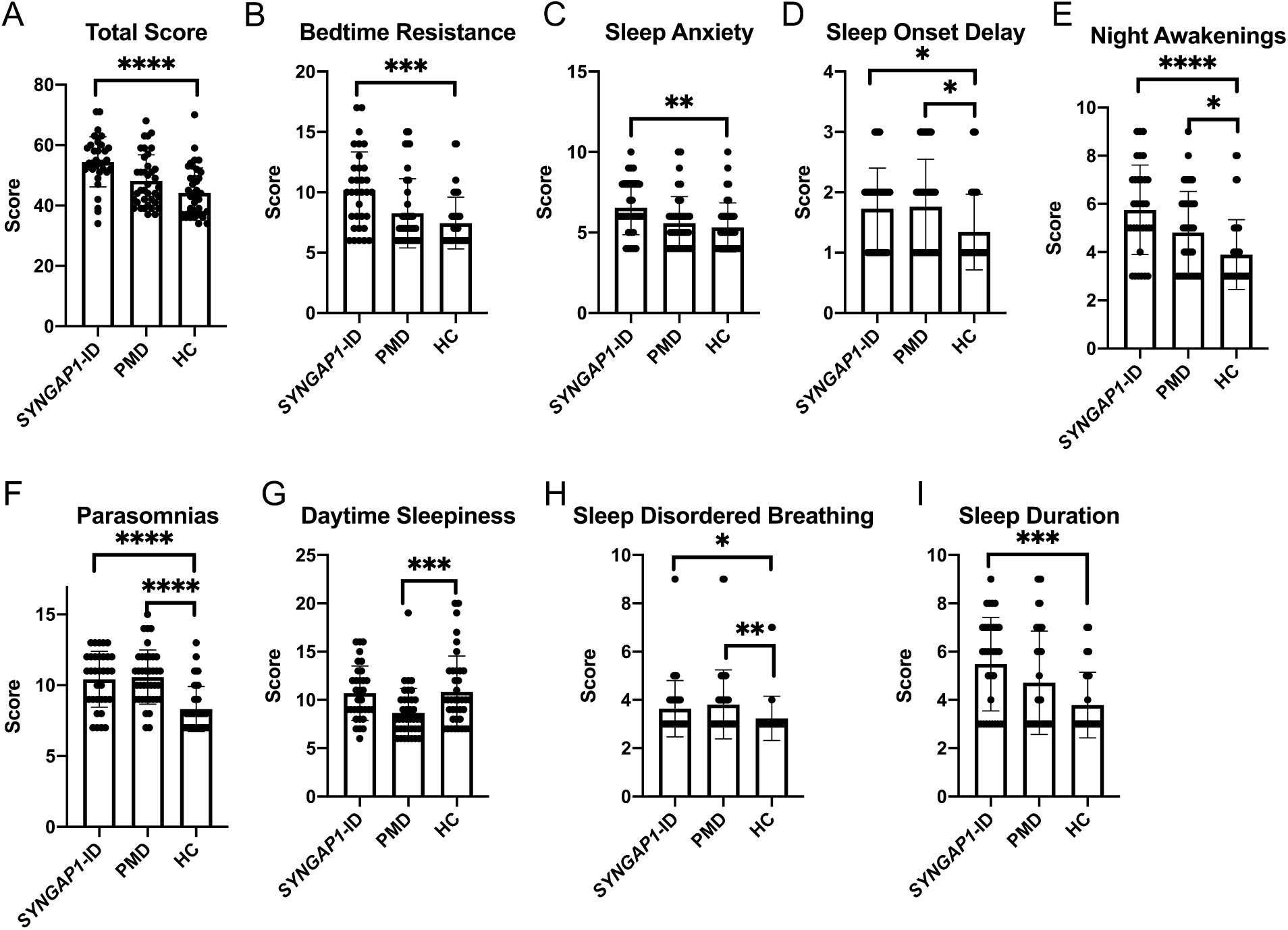
Total (A) and sub-scale scores for *SYNGAP1*-related Intellectual Disability (*SYNGAP1*-ID), Phelan-McDermid syndrome (PMD) and healthy controls (HC). Sub-scales: (B) Bedtime Resistance (C) Sleep Anxiety (D) Sleep Onset Delay (E) Night Awakenings (F) Parasomnias (G) Daytime Sleepiness (H) Sleep Disordered Breathing (I) Sleep Duration. Dunn’s multiple comparisons test *p<0.05, **p<0.01, ***p<0.001,****p<0.0001.

We next parsed our data into those individuals under 10 years of age and those 10 years old or older to determine if the sleep disorders identified in these synaptopathies are age dependent (Figure 2 and Table 2). For those individuals under 10 years old, we again found *SYNGAP1*-ID individuals have a total score (M=54.6±7.0) significantly elevated (p<0.01) compared to healthy controls (M=47.1±8.6). In contrast to the total data, individuals with PMD under 10 years old did not differ (M=47.9±9.6) from healthy controls. The individual sub-scales of bedtime resistance, sleep onset delay, night awakenings and sleep duration were all significantly worse in subjects with *SYNGAP1*-ID than healthy controls but did not significantly differ in PMD. Similar to the data from all ages, subjects with either *SYNGAP-ID* or PMD who were less than 10 years old were significantly more likely to have parasomnias than healthy controls. Overall, these data demonstrate similar abnormalities for the *SYNGAP1*-ID subjects under 10 years of age to the entire *SYNGAP1*-ID population of this study. In contrast, PMD subjects under 10 years of age have fewer significant sleep problems compared with the entire PMD study population.

**Figure 2.**
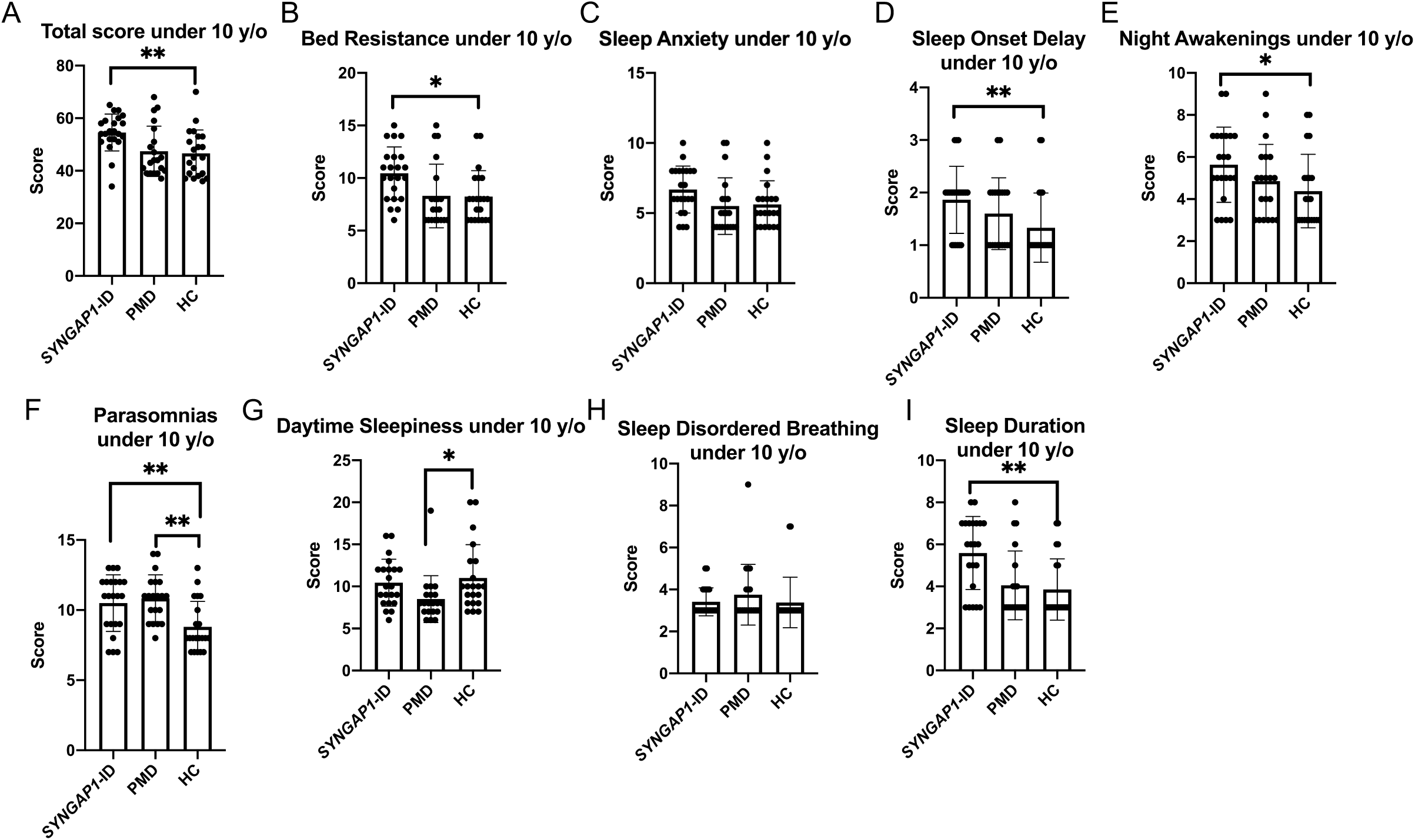
Total (A) and sub-scale scores for *SYNGAP1*-related Intellectual Disability (*SYNGAP1*-ID), Phelan-McDermid syndrome (PMD) and healthy controls (HC) under 10 years of age. Sub-scales: (B) Bedtime Resistance (C) Sleep Anxiety (D) Sleep Onset Delay (E) Night Awakenings (F) Parasomnias (G) Daytime Sleepiness (H) Sleep Disordered Breathing (I) Sleep Duration. Dunn’s multiple comparisons test *p<0.05, **p<0.01, ***p<0.001, ****p<0.0001.

We next evaluated the subjects 10 years of age and over (Figure 3 and Table III). The overall CSHQ score was significantly elevated in both the *SYNGAP1*-ID (M=54.2±10.9, p<0.001) and PMD (M=49.1±7.4, p<0.05) subjects compared with healthy controls (M=41.2±6.3). Only *SYNGAP1*-ID subjects had increased scores in sleep anxiety compared with healthy controls. Subjects diagnosed with *SYNGAP1*-ID or PMD had significantly elevated scores in bedtime resistance, night-time awakenings, parasomnias and sleep disordered breathing. PMD patients had a trend toward increased sleep onset delay (p=0.06) and sleep duration scores (p=0.06) compared with healthy controls. Overall, these data demonstrate similar abnormalities for the *SYNGAP1*-ID subjects under 10 years of age when compared to *SYNGAP1*-ID subjects over 10 years old. In contrast, PMD subjects under 10 years of age have fewer significant sleep phenotypes compared with PMD subjects over 10 years old.

**Figure 3.**
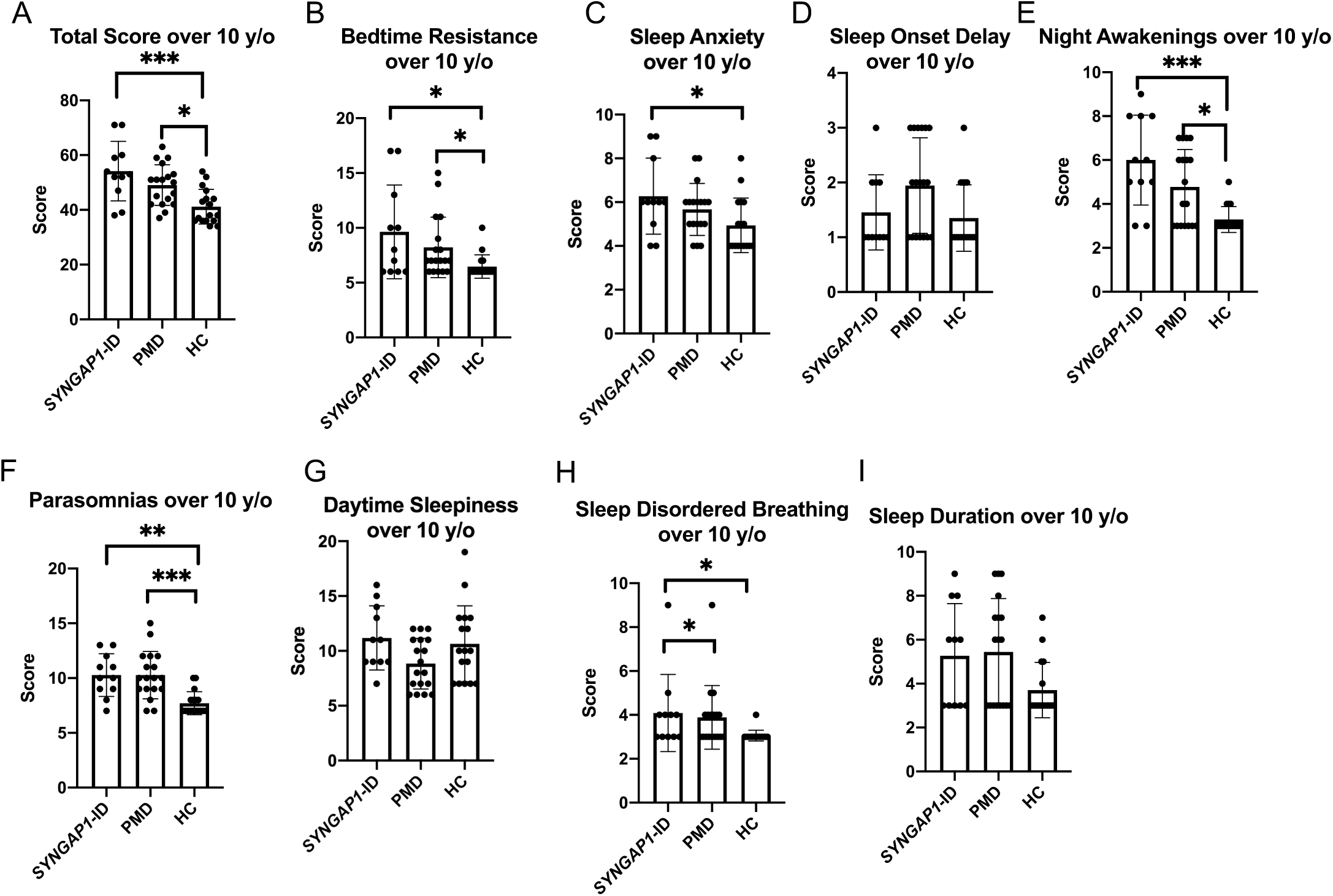
Total (A) and sub-scale scores for *SYNGAP1*-related Intellectual Disability (*SYNGAP1*-ID), Phelan-McDermid syndrome (PMD) and healthy controls (HC) 10 years of age and older. Sub-scales: (B) Bedtime Resistance (C) Sleep Anxiety (D) Sleep Onset Delay (E) Night Awakenings (F) Parasomnias (G) Daytime Sleepiness (H) Sleep Disordered Breathing (I) Sleep Duration. Dunn’s multiple comparisons test *p<0.05, **p<0.01, ***p<0.001, ****p<0.0001.

Recent work has suggested sex bias in phenotypes associated with synaptopathies, particularly in *Syngap1* rodent models^12^. We evaluated our data for a sex bias by directly comparing male and female subjects with each diagnosis (*SYNGAP1*-ID or PMD) as well as our healthy controls. For total scores as well as within each sub-scale, we found no significant difference between male and female scores demonstrating sex does not significantly impact sleep phenotypes in these synaptopathies (data not shown).

Given the striking sleep abnormalities reported by parents of children with *SYNGAP1*-ID, we explored whether abnormalities in sleep architecture were present by polysomnography. We identified four children with pathogenic *SYNGAP1* mutations who had a clinically indicated single night polysomnography recording (Table IV). Each polysomnograph was scored according to the American Academy of Sleep Medicine manual^13^ and reviewed by a board-certified sleep physician. Strikingly for all of these subjects, we identified a reduced amount of time spent in REM sleep (2-17%) than expected (neurotypical 20-25%^14,15^) and increased REM sleep latency of 237-528 minutes (neurotypical 80-150 minutes^14,15^). Total sleep time (410-488 minutes) and sleep efficiency (89-98%) suggest sufficient sleep was recorded. Together, these data demonstrate abnormalities in sleep architecture are associated with *SYNGAP1*-ID.

We also asked parents of our subjects if they were administering medications to improve sleep of their children. We found that 4 out of 39 (10%) of healthy controls took a sleep aid at least occasionally. Three healthy controls used melatonin and one used hydroxyzine. In contrast, 13 out of 25 subjects (52%) with *SYNGAP1*-ID were given at least one sleep aid occasionally or nightly for their sleep disturbance revealing a significant difference from the healthy controls (X^2^=37.88, p<0.0001). The sleep aids used by this population included melatonin, guanfacine, clonidine, diphenhydramine, trazodone, aripiprazole and CBD oil. For the PMD population, 13 out of 39 subjects took at least one sleep aid revealing a similar increase in medication use compared with our healthy controls (X^2^=22.56, p<0.0001). The medications used included melatonin, clonidine, clonazepam, guanfacine, lorazepam, trazodone and zolpidem.

## DISCUSSION

Sleep disturbances have previously been reported in clinical descriptions of synaptopathies including *SYNGAP1*-ID and PMD^4-7^. In this work, we evaluated sleep in these two populations using a standardized, well validated but simple to administer instrument, the Children’s Sleep Habits Questionnaire (CSHQ). We identified significant behavioral abnormalities for both populations including parasomnias, night-time awakenings and sleep onset delay. Overall, sleep scores were significantly worse in *SYNGAP1*-ID subjects than PMD based upon higher total score and more sub-scales of the CSHQ with significant abnormalities in *SYNGAP1*-ID. The unique sleep abnormalities found only in *SYNGAP1*-ID included elevated bedtime resistance, sleep anxiety and worse score for sleep duration.

We parsed out our data based upon age and sex to determine if either is a critical variable to sleep abnormalities in synaptopathies. We chose to parse the data into two groups: those below 10 years of age and those 10 years of age and older to investigate changes in sleep abnormalities in pre-pubertal versus pubertal and post-pubertal individuals in these populations. We found that in general, total score and sub-scale scores were worse for individuals 10 years and over compared with those under 10 years of age. This was most striking for PMD where total CSHQ score was not significantly different in subjects under 10 years of age but significantly elevated in those 10 years old and older compared to healthy controls.

No previous studies have systematically evaluated sleep in *SYNGAP1*-ID. In contrast, one previous study evaluated sleep abnormalities in PMD^9^. Similar to our study, Bro et al. determined that sleep abnormalities are common in individuals with PMD. However, the overall severity as measured by total CSHQ score (M=51.7±9.0) was greater than we observed. The reason for this discrepancy is unclear. Bro et al. did not include healthy controls recruited contemporaneously with the PMD patients, instead relying on historical controls. As such, statistical analysis between PMD and healthy controls was not performed.

Sleep issues are common in NDDs including autism and epilepsy. In particular for epilepsy, it has been hypothesized that abnormal epileptic discharges might disrupt normal sleep architecture leading to sleep disorders^16^. This is potentially most relevant for *SYNGAP1* as >90% of individuals with pathogenic *SYNGAP1* mutations develop epilepsy and have abnormal interictal epileptiform discharges^4,6^. Moreover, epileptiform discharges have been found to be enhanced during sleep in both mice haploinsufficient for the murine orthologue of *SYNGAP1* and patients with pathologic mutations^17^. These enhanced night-time epileptiform discharges might explain the more severe sleep abnormalities discovered in patients with *SYNGAP1* mutations and warrants further investigation.

Intriguingly, we discovered through review of polysomnographs that individuals with *SYNGAP1*-ID demonstrate significantly altered sleep architecture with delayed onset and quantity of REM sleep than expected. These findings occur without particular aberrations in total sleep time or sleep efficiency to suggest common causes of low or delayed REM sleep such as significant wake after sleep onset periods or first-night effects. Furthermore, there were no common features of other significant intrinsic sleep disruptors like sleep-disordered breathing (one patient with significant obstructive sleep apnea had the highest recorded REM sleep percentage), elevated arousal indices, or clear medication effects to account for these findings. A fascinating and discordant finding is the very low sleep latency recorded in these patients (0-7 minutes) in spite of CSHQ scores suggesting they have significant sleep onset complaints. It is possible unaccounted parental measures or medications may have been employed to achieve a successful polysomnograph and low sleep latency, although it does not fully explain the remaining sleep architecture discrepancies. The lower-than-expected REM sleep quantity recorded was observed in conjunction with a greater-than-expected stage N3 sleep in two subjects. Neither feature can be explicitly explained by medication effect. The use of benzodiazepines in (at least) two subjects (2 and 3 from Table IV) may help to explain the low sleep latency in those patients, but we would expect a lower degree of N3 sleep due to benzodiazepines and lamotrigine^18,19^. The three seizures during N3 sleep in one patient may explain the low REM percentage in that one patient only, particularly if the seizures occurred before the first REM period^20^. No systematic approach to evaluating frequency or density of epileptiform discharges was performed in these clinical polysomnographs to evaluate how such discharges may influence sleep architecture, as suggested above. The highly synchronous features of the thalamocortical circuits of N3 sleep and/or paucity of asynchronous REM sleep may support a genetic underpinning to an epileptic network. Future studies with longer recording periods and larger sample size will be critical to determine the consistency of sleep architecture abnormalities in *SYNGAP1*-ID.

Development of targeted treatments for NDDs caused by genetic abnormalities has been slow to be realized. The reason for the difficulty in their development is likely multi-factorial with one cause being lack of quantitative clinical endpoints that are responsive to intervention in the timeline of typical clinical trials. Improvement in sleep represents one potential clinical endpoint for synaptopathies. Studies such as this one can provide the baseline natural history data needed for future clinical trials for these disorders.

## Data Availability

All data will be made available upon reasonable request.

## ACKNOWLEDGEMENTS

We are deeply indebted to all of the subjects and their caregivers that participated in this study. We also are grateful to the patient advocacy foundations: Phelan-McDermid Syndrome Foundation, SynGap Research Fund, Inc. and Bridge the Gap: SYNGAP1 Education and Research Foundation, which aided in patient recruitment. JH and CSH received funding for this study from Bridge the Gap: SYNGAP1 Education and Research Fund. JH also gratefully acknowledges the generous support of the Joan and Stanford Alexander Family Foundation as well as The Robbins Foundation and Mr.Charif Souki.

## Preprint Disclosure

A preprint version of this manuscript appears in medRxiv.

